# Challenges in Institutional Ethical Review Process and Approval for International Multicenter Clinical Studies in Lower and Middle-Income Countries: the case of PARITY Study

**DOI:** 10.1101/2024.03.20.24304598

**Authors:** Eliana Lopez Baron, Qalab Abbas, Paula Caporal, Asya Agulnik, Jonah E. Attebery, Adrian Holloway, Niranjan “Tex” Kissoon, Celia Isabel Mulgado-Aguas, Kokou Amegan-Aho, Marianne Majdalani, Carmen Ocampo, Havugarurema Pascal, Erika Miller, Aimable Kanyamuhunga, Atnafu Mekonnen Tekleab, Tigist Bacha, Sebastian González, Adnan T. Bhutta, Teresa B. Kortz, Srinivas Murthy, Kenneth E. Remy, the Global Health Subgroup of the Pediatric Acute Lung Injury and Sepsis Investigators (PALISI) Network

**Affiliations:** Departamento de pediatría y cuidado crítico Pediátrico. Hospital Pablo Tobón Uribe. Departamento de Pediatría. Universidad de Antioquia. Medellín, Colombia; Department of Pediatrics and Child Health, Aga Khan University Hospital, Karachi, Pakistan; Health Systems Program, Department of International Health, Johns Hopkins Bloomberg School of Public Health, Baltimore, MD, USA; Red Colaborativa Pediátrica de Latinoamérica (LARed Network), La Plata, Buenos Aires, Argentina; Division of Critical Care and Pulmonary Medicine, Department of Pediatrics, St Jude Children’s Research Hospital, Memphis, TN, USA; Department of Global Pediatric Medicine, St Jude Children’s Research Hospital, Memphis, TN, USA; Barrow Global, Department of Neurosurgery, Barrow Neurological Institute, St. Joseph’s Hospital and Medical Center, Phoenix, Arizona, USA; Division of Pediatric Critical Care Medicine, Department of Pediatrics, University of Maryland School of Medicine, Baltimore, MD, United States; Department of Pediatrics, University of British Columbia, Vancouver, BC, Canada; Division of Critical Care, British Columbia Children’s Hospital, Vancouver, BC, Canada; Unidad de Terapia Intensiva Pediátrica, Hospital General León. León, Guanajuato. México; Department of Pediatrics and child health, School of Medicine, University of Health and Allied Sciences, Ho, Ghana; Division of Pediatric Intensive Care Unit, Department of Pediatrics and Adolescent Medicine, American University of Beirut Medical Center, Beirut, Lebanon; Grupo Quirón Salud. Clínica Imbanaco. Cali, Valle del Cauca. Colombia; Department of Pediatrics and Child Health, University Teaching Hospital of Butare, Rwanda; Department of Pediatrics and Child Health, University Teaching Hospital of Kigali, Rwanda; Department of Pediatrics and Child Health, St. Paul’s Hospital Millennium Hospital Medical College, Addis Ababa, Ethiopia; Red Colaborativa Pediátrica de Latinoamérica (LARed Network), Montevideo, Uruguay; Departamento de Pediatría y Unidad de Cuidados Intensivos de Niños del Centro Hospitalario Pereira Rossell, Facultad de Medicina, Universidad de la República, Montevideo, Uruguay; Division of Pediatric Critical Care Medicine, Department of Pediatrics, Indiana University School of Medicine, Indianapolis, IA, United States; Division of Critical Care, Department of Pediatrics, University of California, San Francisco, San Francisco, CA, United States; Institute for Global Health Sciences, UCSF, San Francisco, CA, USA; Department of Anesthesiology, Pharmacology and Therapeutics, University of British Columbia, Vancouver, BC, Canada; Division of Pediatric Critical Care Medicine, Department of Pediatrics, Rainbow Babies and Children’s Hospital, Case Western Reserve University, Cleveland, OH, United States

## Abstract

**Objectives:** To describe the regulatory process, variability and challenges faced by pediatric researchers in low- and middle-income countries (LMICs) during the institutional review board (IRB) process of an international multicenter observational point prevalence study (Global PARITY).

**Design:** A 16-question multiple-choice online survey was sent to site principal investigators (PIs) at PARITY study participating centers to explore characteristics of the IRB process, costs, and barriers to research approval. A shorter survey was employed for sites that expressed interest in participating in Global PARITY and started the approval process, but ultimately did not participate in data collection (non-participating sites) to assess IRB characteristics.

**Subjects:** PIs from the Global PARITY Study

**Interventions:** None.

**Results:** Ninety-one sites pursued local IRB approval and 46 sites obtained IRB approval and completed data collection. Forty-six (100 %) participating centers and 21 (47%) non-participant centers completed the survey. Despite receiving approval from the study’s lead center and being categorized as a minimal risk study, 36 (78%) of the hospitals involved in PARITY study required their own full board review. There was a significant difference between participating and non-participating sites in IRB approval of a waiver consent and in the requirement for a legal review of the protocol. The greatest challenge to research identified by non-participating sites was a lack of research time and the lack of institutional support.

**Conclusions:** Global collaborative research is crucial to increase our understanding of pediatric critical care conditions in hospitals of all resource-levels and IRBs are required to ensure that this research complies with ethical standards. Critical barriers restrict research activities in some resource limiting countries. Increasing the efficiency and accessibility of local IRB review could greatly impact participation of resource limited sites and enrollment of vulnerable populations.

## Introduction

Investigators are increasingly using international multicenter studies to fill important knowledge gaps in pediatric critical illness (1). The process of conducting research in humans needs formal institutional approval with participation of Institutional Review Boards (IRBs) which are tasked to review the study protocol ensuring that the study is performed according to ethical research standards. After this review, IRBs decide whether the study can continue as originally planned or if it needs protocol modifications to ensure protection of research participants (2,3). However, as this process is far from standardized, wide variation of IRB functioning has been previously demonstrated in terms of revision, time to protocol approval, consenting requirements, among others (4,5).

When conducting international studies, the ethical approval process could become one of the main challenges researchers face, particularly in low- and middle-income countries (LMICs) where research culture and resources differ from their high-income country (HICs) counterparts (6). Researchers from LMICs also face additional hurdles prior to IRB submission as they usually are less experienced in submitting a study for IRB review and have less support to help with the administrative process involved (7). As Michelson et al recently demonstrated in an observational pediatric multicenter study involving more than 100 study sites (80% from high resource regions), this heterogeneity in regulatory oversight is a time-consuming process which impacts study participation, eventually provoking trial drop-out of international researchers (8). Moreover, the Covid–19 pandemic has imposed additional challenges on researchers, such as increased site IRB heterogeneity and complexity introduced by some boards and less rigorous standards implemented by others (9). To date, the burden of IRB challenges met by researchers from LMICs remains understudied.

The Global PARITY study (Pediatric Acute cRitical Illness sTudY) was a prospective, observational, multicenter, multinational point prevalence study designed to measure the burden of acute pediatric critical illness in LMICs (10). To better understand IRB related barriers to participating in multinational research studies in LMICs, we used the Global PARITY as a case study, and explored challenges related to the process of submission and conducting research that may have impeded site participation. The primary objective of this study was to evaluate the characteristics of Global PARITY participating site IRBs and compare participating to non-participating site characteristics.

## Methods

### Study Design, Setting and Population

We used the Global PARITY study platform to evaluate the regulatory processes at each participating site. Global PARITY was an unfunded prospective, observational, multicenter, multinational point prevalence study conducted in 46 resource-limited hospitals across North, Central, and South America, Africa, the Middle East; and South Asia. Global PARITY measured the prevalence of pediatric acute critical illness, associated outcomes and resource utilization at four time points throughout one year (July 2021-July 2022). One of the pre-planned secondary studies was the present survey exploring the IRB hurdles encountered during the research process. Participating research sites for this study were recruited via established relationships among physician-led pediatric critical care research networks including the World Federation of Pediatric Intensive & Critical Care Societies (WFPICCS), the Global Health subgroup of the Pediatric Acute Lung Injury and Sepsis Investigators (PALISI) Network (www.palisiglobalhealth.org), Red Colaborativa Pediátrica de Latinoamérica (LARed Network). Global PARITY was coordinated by the Department of Pediatrics at the University of Maryland and has been deemed exempt by the University of Maryland (IRB, HP-00086107). Participating sites were required to obtain local Institutional Review Board (IRB) approval prior to participation.

### Survey Development and Distribution

A subset of Global PARITY investigators and core coordinators with expertise in multinational pediatric critical care research in LMICs developed 16 multiple-choice and categorical questions. The survey was conducted in English and Spanish, according to the sites’ location and was reviewed by researchers before being distributed to site principal investigators. We planned to limit the survey to less than 15 minutes’ completion time. A statement describing survey purpose, length, and the study investigators was included in the introductory page of the survey. A consent statement was included in the introduction of the survey.

After survey development, the Principal Investigator (PI) from each of the 46 participating sites was asked to complete the ethics approval survey after the study’s first two sampling periods, and a follow-up reminder was issued a week later. The survey was a part of a larger investigation on pediatric acute care infrastructure and resource availability, which also included demographic information on the participant sites. An additional shorter survey version was developed for sites that expressed interest in participating in Global PARITY and started the approval process, but ultimately did not participate in data collection (non-participating sites) to assess their IRB characteristics. This shorter survey included a Likert-type scale classifying the barriers for development of research into five levels (not a barrier; somewhat a barrier; neutral; moderate barrier; significant barrier), according to how they felt those barriers might interfere with carrying on research projects. The University of Maryland’s Research Electronic Data Capture (REDCap) tool was used to collect the data from survey responses (11).

### Statistical Analysis

Categorical variables were analyzed using absolute and relative frequencies. Quantitative variables were described, according to their distribution, using means or medians and their respective dispersion measurements (standard deviation, interquartile range or percentiles). Comparison between participant and non-participant sites were done using t-test, Fisher or Chi-square test according to the type and distribution of the variable. Results of the Likert scale were described by calculating the proportion of the institutions within each the level of the barrier. Analysis was done using R version 4.2.1.(17)

## Results

A total of 91 sites pursued local IRB approval. Forty-six sites (50%) were approved and accepted to collect data. The survey was electronically sent to the principal investigators of all centers, with response rates of 46/46 (100%) from participating centers and 21/45 (47%) from non-participant centers for a total of 67 sites. The role of principal investigators was physician in 57% (N=38) of the surveyed centers, and 75% (N= 51) of the institutions were classified by survey respondents as public hospitals with university affiliation. Table 1 shows the characteristics of the institutions and role of the principal investigator in both participating and non-participant institutions.

**Table 1.** Characteristics of participant and non-participant institutions.

|  | *Total (%)*<br>*n= 67* | *Participants*<br>*(%) n= 46* | *Non-participants*<br>*(%) n=21* |
| --- | --- | --- | --- |
| *Role of the principal investigator* |  |  |  |
| Assistant physician | 38 (57) | 22 (48) | 16 (76) |
| Administrator | 8 (12) | 3 (7) | 5 (24) |
| Clinical or medical officer | 8 (12) | 8 (17) | 0 |
| Medical intern | 6 (9) | 6 (13) | 0 |
| Resident | 6 (9) | 6 (13) | 0 |
| Medical student | 1 (1) | 1 (2) | 0 |
| *Type of hospital* |  |  |  |
| Public | 51 (75) | 36 (77) | 15 (71) |
| Private | 14 (21) | 9 (19) | 5 (24) |
| Both | 2 (3) | 1 (2) | 1 (5) |
| Don't know | 1 (1) | 1 (2) | 0 |
| *University affiliation* |  |  |  |
| Yes | 53 (78) | 34 (72) | 19 (90) |
| No | 15 (22) | 13 (28) | 2 (10) |

### Characteristics of IRB process

Table 2 depicts the characteristics of IRB process for both participant and non-participant locations. IRB committees met once every one to two months at 56% (N=37) of sites. The average time needed for a typical research protocol approval was 35 days (range 19.5 - 71 days) for participating sites and 32 days (range 15.2 - 57.8 days) for non-participant sites. There was no difference in the associated cost for the IRB process or mandatory translation into the local language between participant and non-participant sites. Only two non-participant institutions disclosed the values of the costs of the IRB process, so no numerical cost comparisons could be done.

**Table 2.**
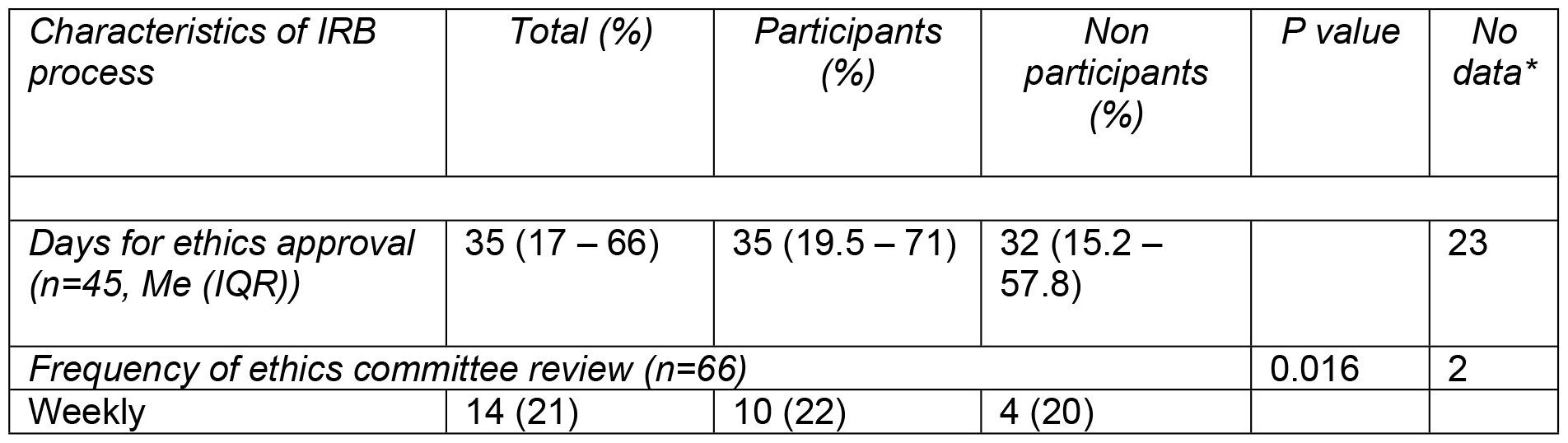
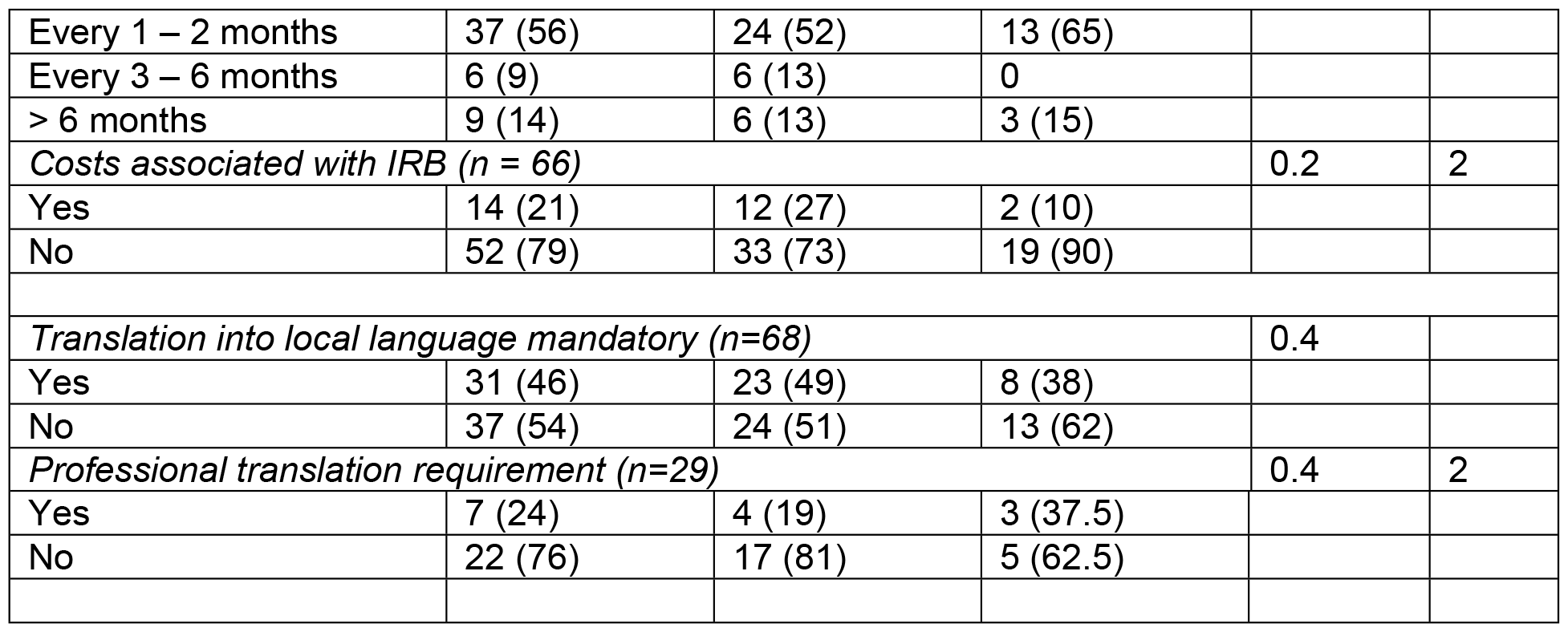
Characteristics of the Ethics Committee / IRB process.

### IRB characteristics

There was no significant difference between participant and non-participant sites in terms of full IRB review requirements for study protocol approval, but there was a significant difference between whether the IRB authorized a waiver of consent (82% vs. 47%, respectively, p=0.015). There was also a significant difference in the requirement for a legal review of the protocol; participating sites required a legal review less frequently than non-participating sites (24% vs. 52%, respectively, p = 0.021). Other IRB requirements did not vary significantly between participating and non-participating sites (Table 3).

**Table 3.** IRB characteristics for all the sites.

| *IRB functioning* | *Total (%)* | *Participants (%)* | *Non participants (%)* | *P value* | *No data\** |
| --- | --- | --- | --- | --- | --- |
| *Full board review to grant approval (n=62)* |  |  |  | 0.5 | 6 |
| Yes | 47 (76) | 36 (78) | 11 (69) |  |  |
| No | 15 (24) | 10 (22) | 5 (31) |  |  |
| *IRB approval of waived consent (n=60)* |  |  |  | 0.015 | 8 |
| Yes | 44 (73) | 37 (82) | 7 (47) |  |  |
| No | 16 (27) | 8 (18) | 8 (53) |  |  |
| *Data Sharing agreement (DTA) requirement (n=65)* |  |  |  | 0.2 | 3 |
| Yes | 15 (23) | 8 (18) | 7 (33) |  |  |
| No | 50 (77) | 36 (82) | 14 (67) |  |  |
| *Approval from a separate institution, without local review (n=68)* |  |  |  | 0.7 |  |
| Yes | 12 (18) | 9 (19) | 3 (14) |  |  |
| No | 56 (82) | 38 (81) | 18 (86) |  |  |
| *Legal review of protocol (n=67)* |  |  |  | 0.021 | 1 |
| Yes | 22 (33) | 11 (24) | 11 (52) |  |  |
| No | 45 (67) | 35 (76) | 10 (48) |  |  |

\* Different sample sizes are found for some variables due to missing data,

### Characteristics of the IRB in non-participant sites

Table 4 provides a detailed analysis of the characteristics, barriers, and opportunities to participate in research for non-participating sites. Seventy-one percent of the non-participating sites (N=15) reported having previously participated in multicenter research studies; however 95% (N = 19) reported not having protected time, 71% (N=15) not receiving institutional support for research. Furthermore, 63% (N=14) indicated a lack of research-trained investigators (epidemiologist, statistics). Sixty-two percent (n=13) of those questioned about the advantages of undertaking research for academic or institutional purposes did not mention any advantages.

**Table 4.** Characteristics of the research process in non-participating sites.

|  | *Non participants (%)*<br>*n=21* |
| --- | --- |
| *Benefit/advantage of doing research\** |  |
| Economic | 2 (10) |
| Time for research | 2 (10) |
| Academic | 9 (43) |
| None | 13 (62) |
| *Protected time to research* |  |
| Yes | 1 (5) |
| No | 19 (95) |
| *Availability of Support for research (epidemiologist, writing manuscripts, translation)* |  |
| Yes | 6 (29) |
| No | 15 (71) |
| *Previous participation in research* |  |
| Yes | 15 (71) |
| No | 6 (29) |
| *Previous participation in multicenter multinational research* |  |
| Yes | 13 (62) |
| No | 8 (38) |
| *If so, did this result in a publication?* |  |
| Yes | 14 (88) |
| No | 1 (12) |
| *Additional research training* |  |
| Yes | 7 (33) |
| No | 14 (63) |

\*More than one option could be selected \*\*One institution did not report data

The barriers to research participation, as indicated by the non-participating sites, are depicted in Figure 1. The greatest challenge was a lack of research time, which 57% of respondents rated as a moderate to major obstacle, and the lack of institutional support. The absence of institutional funding for research, the challenge of collecting data, and the availability of staff to help with data collecting were other significant obstacles. Legal restrictions and access to technology for research collaboration were not seen as significant barriers.

**Figure 1.** Barrier levels for research participation among non-participant institutions.

## Discussion

This study evaluated characteristics of the Institutional Review Board (IRB) process in low- and middle-income countries (LMICs) in the context of approving the Global PARITY, a multicenter minimal risk study. We identified significant barriers to participating in international multicenter research studies. These findings represent an important first step to help overcome these barriers and to increase the participation of resource-limited centers in future studies, and thus improve the outcomes in resource limited settings.

One key observation was that, despite receiving approval from the study’s lead center and being categorized as a minimal risk study, most of the hospitals involved in this study required their own full board review. This result is consistent with earlier international research projects, including the sepsis prevalence, outcomes, and therapies study (SPROUT), wich demonstrated comparable findings in resource-limited settings (8) regarding barriers and challenges for IRB approval. These barriers could contribute to the difference in sepsis outcomes between institutions from HICs and LMICs. Global collaborative research is crucial for enhancing our understanding of relevant critical pediatric care conditions like sepsis (12) and pediatric acute respiratory distress syndrome (13), in hospitals of all resource-levels (14). The process of evaluating approaches, resources and outcomes in resource limited centers is a first step in the elaboration of strategies to improve outcomes and reduce disparities. The differences of population characteristics, prevalence of diseases and centers’ resources and the proposal of strategies suited to local setting, make this research relevant for populations in low-resource settings (15).

Our study examined the difficulties experienced by non-participating sites, identifying time limits, funding shortages, and a shortage of research workers as major obstacles, combined with a considerable lack of institutional support (62%) and protected institutional time for research. Another important finding was the absence of formal research training, which represents a barrier to research, especially to analyze local data with the aim to evaluate the performance in resource limited populations.

The Institutional Review Board (IRB) is a crucial component of research because it protects study participants and maintains ethical standards (2). However, obstacles encountered with regional IRBs or ethics committees have limited the involvement of some sites in multicenter trials, thus limiting the benefits of such research to patients in those settings. These difficulties make it less likely for centers to participate in multicenter research, which involves gathering data as well as attending meetings and working together on collaborative writing projects. Furthermore, language barriers may provide serious challenges for international research collaborations (6,16).

The strengths of this study include its focus on countries with limited resources, where exploring critical care conditions in children is especially valuable. These conditions may be influenced by resource availability, geographical location, and other contextual elements. To encourage greater participation of resource-limited centers, it may be beneficial to establish a simpler or standardized research involvement process and address language barriers. Centralized IRBs could also streamline the approval process.

This study has several limitations. Although the Global PARITY was international, there was overrepresentation of centers in Latin America (43%) (10). Our findings of barriers to research may not fully reflect conditions in other resource-limited settings with different characteristics, such as language and time frame. Additionally, the Global PARITY study was a point prevalence study, and our findings may not describe all barriers faced in other types of study designs. Further research could explore additional barriers and potential solutions in other study types and settings.

## Conclusion

Global collaborative research is essential, and IRBs are critical to ensure that this research complies with ethical standards, but the benefits of this kind of research may be constrained by obstacles to IRB approval. Critical barriers to study site participation were absence of institutional support for research, which coexisted with staffing shortages, restricted protected research time, financial assistance, and inadequate training, which are modifiable factors. These barriers restrict research activities in some resource limiting countries. Increasing the efficiency and accessibility of local IRB review could greatly impact participation of resource limited sites and enrollment of vulnerable populations.

## Data Availability

All relevant data are within the manuscript

## Acknowledgments

The authors thank to Global PARITY study collaborators for help survey data collection. *Argentina:* Guillermo Kohn Loncarica (Hospital Profesor Dr. Juan P. Garrahan), Sofia Esposto (Hospital Sor Maria Ludovica); Aurora Leonor Pedroza (Hospital Público Materno Infantil); Curi, Claudia Patricia (Hospital de niños Santísima Trinidad), Marina Giulietti (Hospital San Roque de Gonnet); *Barbados:* Kandamaran Krishnamurthy (Queen Elizabeth Hospital); *Brasil:* Adriana Teixeira Rodrigues (Hospital das Clinicas); *Colombia:* Carmen E. Ocampo.(Clínica Imbanaco); Mayerly Milena Palencia Bocarejo (Hospital Simon Bolivar), Isabel Cristina Monje Cardona (Hospital de Suba), Lucia Carolina Hernandez Somerson (Hospital de Engativá), Nayibe Hincapie (Hospital General de Medellin); Carmen Rossy Ramirez Hernandez (Hospital María Inmaculada); Nataly Ávila Guerrero (Clínica El Rosario), Freddy Israel Pantoja Chamorro (Hospital Infantil Los Ángeles); Liliana Patricia Jurado Salcedo (Clínica Infantil Santa María del Lago); *Mali:* Adama Mamby Keita (University Hopital Gabriel Toure); *Ethiopia:* Emnet Tesfaye (Hawassa University Comprehensive Specialized Hospital); Atnafu Mekonnen Tekleab (St Paul’s Hospital Millennium Medical College) *Ghana:* Alhassan Abdul-Mumin (Tamale Teaching Hospital); Edna Okaikor Obodai (Cape Coast Teaching Hospital); Rita Fosu Yeboah (Ashanti Regional Hospital); Gustav Kumu Nettey (Komfo Anokye Teaching Hospital), Charlyne Kilba (Greater Accra Regional Hospital), Jacqueline Gyapomaa Asibey (Holy Family Hospital Techiman), Afua Kwakyewaa Osew-Gyamfi (Holy Family Hospital Techiman), Afua Kwakyewaa Osew-Gyamfi (Eastern Regional Hospital), Kokou H. Amegan-Aho (Ho Teaching Hospital); *India:* Sheetal Agarwal (AIIMS), *Kenya:* Amelie von Saint A. and Arianna Shirk (Aic kijabe hospital); *Mexico:* Celia Isabel Mulgado Aguas (Hospital General Leon); *Lebanon:* Amal Rahi (American University of Beirut Medical Center); *Mongolia:* Solongo Orosoo (National center for maternal and child health); *Nigeria:* Tagbo Oguonu (University of Nigeria Teaching Hospital Ituku/Ozalla), Halima Kabir (Aminu Kano Teaching Hospital/Bayero University) *Pakistan:* Qalab Abbas (Aga Khan University Hospital); Arif Fehmina (Dr. Ruth KM Pfau Civil Hospital Karachi); Muhammad Irfan Habib (SMBBMU Childlife Foundation Larkana); *Rwanda:* Christian Umuhoza and Aimable Kanyamuhunga (Centre Hospitalier Universitaire de Kigali), Pascal Havugarurema (Centre hospitalier universitaire de butare); *Peru:* Jesús Angel Domínguez Dominguez (Hospital Nacional Edgardo Rebagliati Martins); *Tanzania:* Raya Yusuph. (Muhimbili National Hospital); *Turkey:* Çağlar Ödek (Bursa Uludağ University Hospital) *Uganda:*Tagoola Abner and Tenywa Emmanuel. (Jinja Regional Referral Hospital) *Uruguay:* Alberto Serra (Casa de Galicia), Javier Prego (Hospital Pereira Rossell)

